# Anterior Connectivity Critical for Recovery of Connected Speech after Stroke

**DOI:** 10.1101/2022.02.04.22270450

**Authors:** Junhua Ding, Tatiana T. Schnur

## Abstract

Connected speech recovers to different degrees across people after left hemisphere stroke, but white matter predictors of differential recovery from the acute stage of stroke are unknown. We assessed changes in lexical-syntactic aspects of connected speech in a longitudinal analysis of 40 individuals (18 females) from the acute stage of left hemisphere stroke (within an average 4 days post-stroke) to subacute (within 2 months) and chronic stages (early: 6 months, late: 1 year) while measuring the extent of acute lesions on white matter tracts to identify tracts predictive of recovery. We found that acute damage to the frontal aslant tract led to decreased recovery of the fluency and structural complexity of connected speech during the year following left hemisphere stroke. Results were independent of baseline performance, overall lesion volume, and the proportion damage to tract-adjacent gray matter. This longitudinal analysis from acute to chronic stroke provides the first evidence that recovery of fluent and structurally complex spontaneous connected speech requires intact left frontal connectivity via the frontal aslant tract. That the frontal aslant tract was critical for recovery at early as well as later stages of stroke demonstrates that anterior connectivity plays a lasting and important role for the reorganization of function related to the successful production of connected speech.

## Introduction

Left-hemisphere stroke is a high-frequency and high-risk disease that profoundly affects communication by impairing the ability to produce words^1,2^ and combine them into connected speech.^3–6^ To facilitate language intervention, it is important to identify factors that predict degree of connected speech recovery soon after stroke. Damage to disparate brain regions and their connections impairs word retrieval and its recovery,^7–9^ but the neural predictors of the recovery of connected speech are unknown. In this large-scale longitudinal analysis (n=40), we examined the role of white matter tract integrity in the recovery of connected speech after acute left hemisphere brain damage during the first year after stroke.

Successfully producing connected speech requires retrieving words and combining them into phrases and sentences while avoiding long hesitations. Connected speech is impaired to different degrees after stroke as a result of damage to disparate brain regions in the left hemisphere. Anteriorly, damage to the left inferior frontal gyrus (IFG) impairs the ability to produce syntactically well-formed sentences and to retrieve grammatically marked words (pars opercularis;^5^ cf.^10^) and their verbal quantity (i.e. number of total or distinct content words) and quality (i.e. content words/total words; pars opercularis;^3^ cf.^11^). Posteriorly, the posterior superior/middle temporal gyri and temporoparietal junction (angular gyrus and supramarginal gyrus) are also crucial regions for connected speech. Damage negatively impacts sentence complexity and the retrieval of nouns (posterior superior temporal sulcus and angular gyrus;^5^ cf.^10^). In addition, a reduction in distinct words to total words produced (type/token ratio; posterior superior/middle temporal gyri, angular gyrus and supramarginal gyrus^4,6^) and increased semantic and unrelated word errors (posterior superior/middle temporal gyri and angular gyrus^12^) are also associated with damage to the temporal pole and anterior portions of all temporal gyri. Together, a network of anterior to posterior brain regions appears critical for the distinct abilities required to successfully produce connected speech.

The degree to which regions are connected also contributes to connected speech performance and may, at least as seen with general cognitive abilities, better account for impairments in comparison to lesion topography^13–15^ (cf.^11^). One well-defined pathway important for connected speech is the arcuate fasciculus (AF). Most studies have focused on the relation between connected speech impairment and damage to the long segment of the AF (LSAF), which connects the posterior part of the pars opercularis of the IFG (BA 44) with middle and inferior temporal gyri.^16,17^ Its damage in chronic stroke patients is associated with impairments in different aspects of word retrieval during connected speech including the speed with which overall words and narratively relevant words are produced (i.e. fluency^18–21^). Similar impairments in connected speech are also seen after chronic damage to other parts of the AF. For example, damage to the anterior segment of the AF (ASAF), connecting the posterior part of the pars opercularis of the IFG with the supramarginal gyrus is also associated with impairments in the fluency as well as a general composite measure of connected speech (a combination of appropriate information, fluency, syntactic variety and accuracy measures^11,22^). After accounting for the contribution of the ASAF, chronic damage to another tract, the uncinate fasciculus (UF) which connects the IFG (pars orbitalis; BA 47) with the anterior temporal lobe (BAs 20 and 38^23^) was also associated with speech fluency impairments.^22^ Thus, white matter pathways which connect the IFG with other cortical areas play a critical role for lexically-based aspects of connected speech.

However, not all IFG associated tracts are involved in connected speech production. A less-explored tract, the frontal aslant tract (FAT) which connects the pars opercularis of the IFG with the anterior supplementary and pre-supplementary motor areas^24,25^ has not been directly found to impact either lexical or discourse aspects of connected speech.^11,21^ We are aware of one study where lesion damage associated with the reduced number, diversity, and speed of content words produced during picture descriptions and procedural discourse, overlapped with atlas-defined FAT (along with the LSAF and the ASAF) in a group of 46 speakers with chronic aphasia.^3^ These results point to a potential role for the FAT in lexical retrieval and fluency, but statistical evidence of this tract’s function is needed. Similarly, there is no evidence of which we are aware of a role in connected speech for the IFOF^21,22^ which connects the IFG (BAs 44, 45 and 47) with the superior anterior temporal lobe and posterior temporal lobe.^26,27^ Lastly, with regards to pathways which connect posterior regions, chronic damage to the posterior segment of the AF (PSAF) which connects the posterior middle temporal gyrus with the angular gyrus was not associated with speech fluency of connected speech^22^ (cf. ^21^ for evidence regarding how ventral tract damage to the ILF which connects the anterior temporal lobe with the posterior temporal lobe affects discourse). Thus, the evidence to date suggests that chronic impairments in different aspects of word retrieval during connected speech are associated with damage to tracts which connect the IFG with other regions (LSAF, ASAF, UF) although this relationship does not extend to all anteriorly connected tracts (FAT, IFOF).

It is unclear why damage to white matter tracts that connect different parts of the language network associated with different language functions have not been strongly differentiated with regards to connected speech. However, because study enrollment is usually restricted to those participants with clinically diagnosed aphasia, the severity of language impairments was likely accompanied by more extensive brain damage. As a result, participants may have damage to multiple brain regions simultaneously, thus equally impacting different white matter tracts. Second, some studies used measures combining multiple aspects of connected speech (e.g., Western Aphasia Battery-speech fluency^22^) and thus it is unknown which aspect of connected speech was most strongly related to a particular segment. Lastly, it is possible that white matter tracts become differentiated with regards to language function to different degrees across individuals because function is reorganized by the chronic stroke stage.

In the only study we are aware of reflecting functional reorganization of connected speech after stroke, Keser et al.^21^ examined the relationship between the degree of early (within < 3 months after stroke) damage to white matter connectivity and recovery of connected speech in ten speakers after stroke. No results were significant in the longitudinal analysis after multiple comparison correction. However, before applying correction, the integrity (i.e. radial diffusivity) of the left LSAF within < 3 months after stroke was correlated with lexical-semantic retrieval/informativeness (i.e. the number of content units produced during picture description) during the chronic phase (4-13 months after stroke). This result is similar to results in participants with chronic stroke.^3,18–20^ Regarding how early white matter tract damage affects the production of syntactic aspects of connected speech, significant results were obtained only when examining the relationship between early damage and early performance. Damage to the left ILF (i.e. fractional anisotropy) predicted a measure commonly used to examine discourse, that is, the cohesiveness of one sentence to another measured by the use of personal pronouns to refer to previously named agents. However, coherence reflects not only syntax, but also lexical, semantic and executive control abilities^28,29^ making it difficult to interpret which aspect of connected speech critically depended on the ILF. These results highlight the potential importance of the AF and ILF for the recovery of broad aspects of connected speech, but the small subject sample size and lack of lexical-syntactic connected speech measures in participants during the acute stage of stroke leave open the question of which white matter tracts are critical for the recovery of connected speech.

### Current Study

Our aim was to predict the recovery of connected speech using the degree to which white matter tracts were damaged within 1-4 days after left hemisphere stroke. To our knowledge there is no evidence to date concerning how white matter tract damage at the acute stage of stroke affects recovery of abilities to combine words into phrases and sentences, both critical components of successfully produced connected speech. Our study offers distinct advances over previous work. First, we used an ecologically relevant measure of connected speech, spontaneous narrative story-telling. Second, we hand-extracted continuous measures of lexical-syntactic connected speech abilities using quantitative production analysis (QPA^30,31^) which reflects more precise measurements of language abilities in comparison to often used qualitative scoring. Third, we examined the integrity of white matter tracts in participants identified with radiological signs of left hemisphere acute stroke independent of language deficit severity which increased variability in lesion size and location.^5^ In contrast to speakers with chronic aphasia who typically have large lesions spanning adjacent cortical regions,^32–34^ by including participants with smaller lesions we could disentangle the contribution to function of differentially damaged white matter tracts while also minimizing effects of overall lesion volume. We intentionally focused on white matter tracts which connect gray matter regions previously found to be involved in language production and also additional segments of the AF (i.e. ASAF and PSAF). Fourth, because acute, subacute and chronic phases of stroke display dynamic brain reorganization mechanisms which recruit different brain regions,^35–37^ we assessed longitudinal behavior acutely (< 4 days after stroke) and at least one to three additional time points during the year after stroke (subacute: 2 months; early chronic: 6 months; late chronic: 12 months).

We predicted white matter tract involvement in recovery of different aspects of connected speech based on previous evidence of the functional necessity of associated gray matter regions. We expected that recovery of fluency and syntactic accuracy would be influenced by the integrity of anterior connectivity between the IFG (pars opercularis) and supplemental motor (FAT), supramarginal gyrus (ASAF) and middle temporal regions (LSAF).^5,11,18–22^ We expected the recovery of word retrieval and structural complexity would be influenced by the integrity of posterior connectivity (PSAF and ILF) because these functions are associated with the temporoparietal junction or posterior superior/middle temporal gyri damage.^5^ By examining longitudinal recovery in large sample sizes (> 20 participants; total n = 40) at four time points while controlling for baseline performance,^38,39^ we tested for specific white matter mediated mechanisms of recovery from before significant reorganization of brain behavior occurred through subsequent stages of functional reorganization.

## Methods

### Participants

Forty left hemisphere stroke patients (22 male; 35 right-handed; 1 hemorrhagic stroke; age: M = 61, S.D. = 13, range = 20-85 years; education: M = 14, S.D. = 3, range = 8-23 years) were consecutively recruited and tested during the acute phase of stroke (interval between stroke onset and testing: M = 4 days, S.D. = 2, range = 1-12) and follow-up stages post-stroke (subacute: n = 33, interval: M = 56 days, S.D. = 29, range = 23-124; post 6 months: n = 33; interval: M = 204 days, S.D. = 31, range = 163-294; 12 months: n = 27, interval: M = 398 days, S.D. = 49, range = 339-510) from the Memorial Hermann, Houston Methodist and St. Luke’s hospitals’ comprehensive stroke centers in Houston, Texas, USA as part of an ongoing longitudinal project.^5,31,40,41^ Twenty-two participants were tested at all four time points. Subjects met the following inclusion criteria: Native English speaker; No concomitant neurological/psychiatric diseases (e.g., tumor, dementia, epilepsy or depression); No severe visual or auditory deficits. Two patients were recruited with neurological signs of acute left stroke, but no clear lesion was identified from neuroimaging. Three patients had chronic left lesions (> 15 mm^3^;^42^ lesion locations: cerebellum, basal ganglia and medial parietal lobe). To note, we included < 10% of patients with prior chronic stroke because our primary aim was to understand the recovery mechanisms after acute stroke damage and with these limited subject inclusions, prior stroke history is not a significant factor accounting for language recovery.^43^ The control group consisted of 13 non-brain damaged participants (3 male, 11 right-handed) with normal cognitive ability (Mini-Mental State Examination Scores > 26)^44^ matched in age and education with the patient group (|t|’s < 1.74; P’s > 0.09). Mean age and education were 55 (S.D. = 14, range = 37–78) and 16 (S.D. = 3; range = 12–22) years, respectively. Informed consent was approved by the Baylor College of Medicine Institutional Review Board.

### Connected speech assessment

Participants viewed a picture book of the Cinderella story ^45^ for as long as they wished with printed text occluded, and then told the story in their own words without viewing the book.^5,40^ Connected speech narratives were transcribed and scored according to the procedures of QPA^30,46,47^ to yield 13 lexical-syntactic measures of connected speech. The transcription and QPA inter-reliability scoring was originally reported in a previous study.^40^ Briefly summarized, for transcription, two raters scored the middle 30 sec of narratives and for QPA scoring, 10 randomly selected utterances from each of 15 participants who produced more than 10 utterances. For transcriptions, on average across samples, raters achieved high agreement on the number (95%) and identity (93%) of narrative words as well as the segmentation of narrative words into utterances (98%). Across QPA measures, inter-rater reliability was an average 93% (range: 79%-100%).

We used the principal component analysis (PCA) coefficients generated for four connected speech components from the large acute left hemisphere stroke sample (n = 65) in Ding et al.^5^ to calculate component scores for each participant. We used results from the Ding et al.^5^ PCA because its large sample size provides a more reliable estimate of principle component scores than those derived from the current more modestly-sized subject sample. We first z-scored the QPA measure scores relative to control performance because the previous PCA was based on z-scores of the QPA measures. Since the PCA was originally conducted in patients, scaling indicated the performance relative to patients. Therefore, we standardized PCA scores based on the control cohort’s PCA component scores for an intuitive interpretation of impairment degree and direct comparison between components. The four component scores reflected structural complexity, lexical selection, syntactic accuracy and fluency aspects of connected speech. As described in detail by Ding et al.^5^ structural complexity reflected the degree of phrase elaboration, number of sentence embeddings, and sentence length. Lexical selection reflected the ability to produce nouns in comparison to verbs, pronouns, and closed-class words. Syntactic accuracy reflected the ability to produce syntactically accurate speech, including more well-formed sentences, more words within as opposed to outside of sentences and increased production of required determiners. Fluency reflected the number of narrative words produced per minute. For the lexical selection component, we multiplied scores by −1 so that lower scores reflect increased impairment. The test-retest reliabilities for the four component scores were estimated in a subset of 14 participants who performed within control level (> −1.67 S.D. of controls) at 6 months after stroke. The correlations between their performance at 6 and 12 months were calculated. Performance significantly correlated across timepoints for component scores representing structural complexity (r = 0.66, *P* = 0.01), lexical selection (r = 0.59, *P* = 0.03) and fluency (r = 0.73, *P* = 0.003). Syntactic accuracy did not reach statistical significance (r = −0.27, *P* = 0.36), likely due to variability in individual syntactic accuracy recovery during later stages of stroke. Three patients were removed from analyses because their acute and/or follow-up z-scores of the syntax component were extreme outliers (< 25% quartile – 3*interquartile range or > 75% quartile + 3*interquartile range). Therefore, the final patient participant sample sizes across the subacute, post 6 and 12 months groups were 31, 31 and 25 participants, respectively. The final sample size of participants tested at all the time points was 20. We defined the dependent measure of patients’ recovery as the difference between component z-scores at acute and each follow-up stage.

### Neuroimaging acquisition and processing

We acquired diffusion weighted and high-resolution structural scans (T1 and T2 FLAIR) along the axial direction as part of the clinical protocols for admitted acute stroke cases (interval between stroke onset and scan: M = 2 days from stroke onset; S.D. = 2; range = 0-10). The voxel sizes of diffusion-weighted and structural images were 1*1*4.5 mm, and 0.5 * 0.5 * 4.5 mm, respectively.

Acute lesions were delineated manually. We first co-registered the diffusion weighted images with the high-resolution structural images (T1 or T2) using AFNI (https://afni.nimh.nih.gov/).^48^ Lesions were delineated on the diffusion weighted images, using ITK-snap (http://www.itksnap.org/pmwiki/pmwiki.php).^49^ Next, we normalized the individual structural images to the Colin-27 template/MNI space using ANTs registration (http://stnava.github.io/ANTs/).^50^ Finally, we used the affine parameter and diffeomorphic map from the last step to transform individual masks to the MNI space. Due to MRI contraindication, one patient received a CT scan. This lesion was directly delineated on the Colin 27 template based on the CT image.

To quantify the damage to white matter tracts at the acute stage of stroke, we extracted the intersection volume between acute lesion masks and seven primary tracts. Ventral pathways included the IFOF, ILF and UF and dorsal pathways included the FAT and AF (see Figure 1). With regards to the AF, it was further separated into anterior, long, and posterior segments.^51–53^ We defined the tracts using a probabilistic atlas where voxels within white matter tracts were defined using a 50% probability criterion.^25^ Tracts were considered damaged if the intersection volume was above 100 mm^3^.

**Figure 1.**
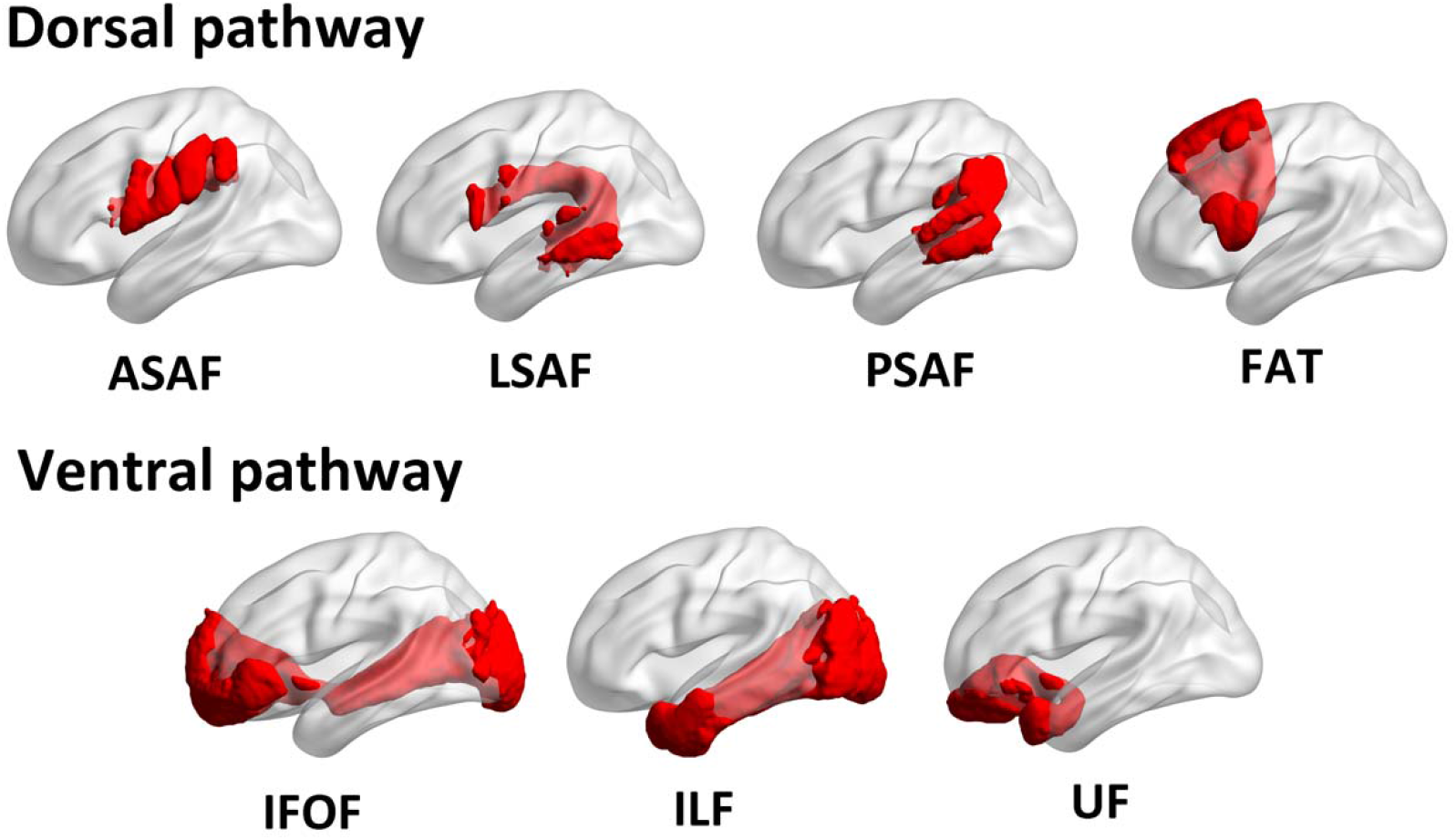
The white matter tracts of interest. ASAF/LSAF/PSAF: anterior/long/posterior segment of arcuate fasciculus; FAT: frontal aslant tract; IFOF: inferior fronto-occipital fasciculus; ILF: inferior longitudinal fasciculus; UF: uncinate fasciculus.

### Statistical analysis

To determine the relationship between white matter tracts and connected speech recovery, we conducted a series of lesion-symptom mapping analyses. Before executing the lesion-symptom mapping analyses, we regressed out participant acute performance (baseline) and total lesion volume from the recovery score.^54,55^ We controlled for acute performance because it was highly correlated with degree of recovery (see Results). Then using LESYMAP (https://dorianps.github.io/LESYMAP/),^56^ we conducted a Brunner-Munzel test to compare the recovery scores between tract-preserved and damaged groups where at least > 10% of individuals had damage to a given tract.^1,57^ To control for multiple comparisons, we applied a Bonferroni correction (corrected *P* = 0.05/7). Then, to confirm the specificity of acute white matter tract damage results independent of adjacently located gray matter damage, we replicated results while controlling for the degree of gray matter damage at tract origins and termini. We defined the gray matter damage as the intersection volume between acute lesion masks and gray matter ROIs extracted from the AAL atlas.^58^ Lastly, we replicated results on the subset of participants tested at all time points. By doing this, we removed the potential influence of individual differences across the three main comparisons comprising overlapping but not identical groups of patients (acute vs subacute/6/12 months).

## Data availability

The data that support the findings of this study are available from the corresponding author, upon reasonable request.

## Results

### Connected speech recovery from the acute stage of stroke

Fluency of connected speech improved to within neurotypical levels by the subacute timepoint for over 50% of participants who had impaired baseline acute performance (< −1.67 S.D. of controls). However, fluency deficits continued to be a problem at chronic stage timepoints for 39-50% of participants who were impaired acutely. In contrast, more of the acutely impaired patients recovered to a neurotypical level in their syntactic impairment during connected speech production (80-100% of acutely impaired patients). Regarding lexical ability, 80% of acutely impaired patients recovered to within neurotypical levels by the subacute time point, and the proportion of participants who recovered from acute impairment remained relatively constant at the 6 and 12 months time points (67–80%, respectively). For structural complexity of connected speech, at the subacute stage, only 67% of acutely impaired patients recovered to within neurotypical levels, but the proportion of acutely impaired patients performing within the neurotypical range rose to 80% 6 months post stroke, and eventually 100% a year after stroke (see Figure 2, Figure 3 and Table 1). When considering those participants who made large gains (recovery > 1 S.D.) but not to within the control range (see medium recovery group; SI Figure 1), the recovery patterns were relatively consistent with the group of participants who recovered to within control levels. Specifically, syntactic accuracy, structural complexity and lexical selection patterns of recovery were similar. However, for fluency, when including participants who made large gains but outside the control range, 70% of acutely impaired participants improved during the year after stroke (in comparison to 50% of acutely impaired participants who improved to within the control range). In the subset of participants who returned at every time point, patterns of connected speech recovery were similar to the larger overlapping subject groups returning across timepoints (see SI Table 1, SI Figure 2 and SI Figure 3). In sum, recovery to neurotypical levels of performance for fluency averaged to approximately one in every two patients who were impaired acutely, while syntactic, lexical selection and structural complexity eventually recovered to neurotypical levels in > 80% patients impaired acutely.

**Table 1.** The number and proportion of patients who recovered to within normal levels (z > − 1.67) after beginning with impairment acutely (z < −1.67) in four aspects of connected speech across three time points in the year following stroke.

|  | Fluency | Syntactic accuracy | Lexical selection | Structural complexity |
| --- | --- | --- | --- | --- |
| Subacute (n = 31) | 10/18 (56%) | 4/5 (80%) | 4/5 (80%) | 2/3 (67%) |
| 6mo (n = 31) | 11/18 (61%) | 6/7 (86%) | 6/9 (67%) | 4/5 (80%) |
| 12mo (n = 25) | 6/12 (50%) | 3/3 (100%) | 4/5 (80%) | 3/3 (100%) |

**Figure 2.**
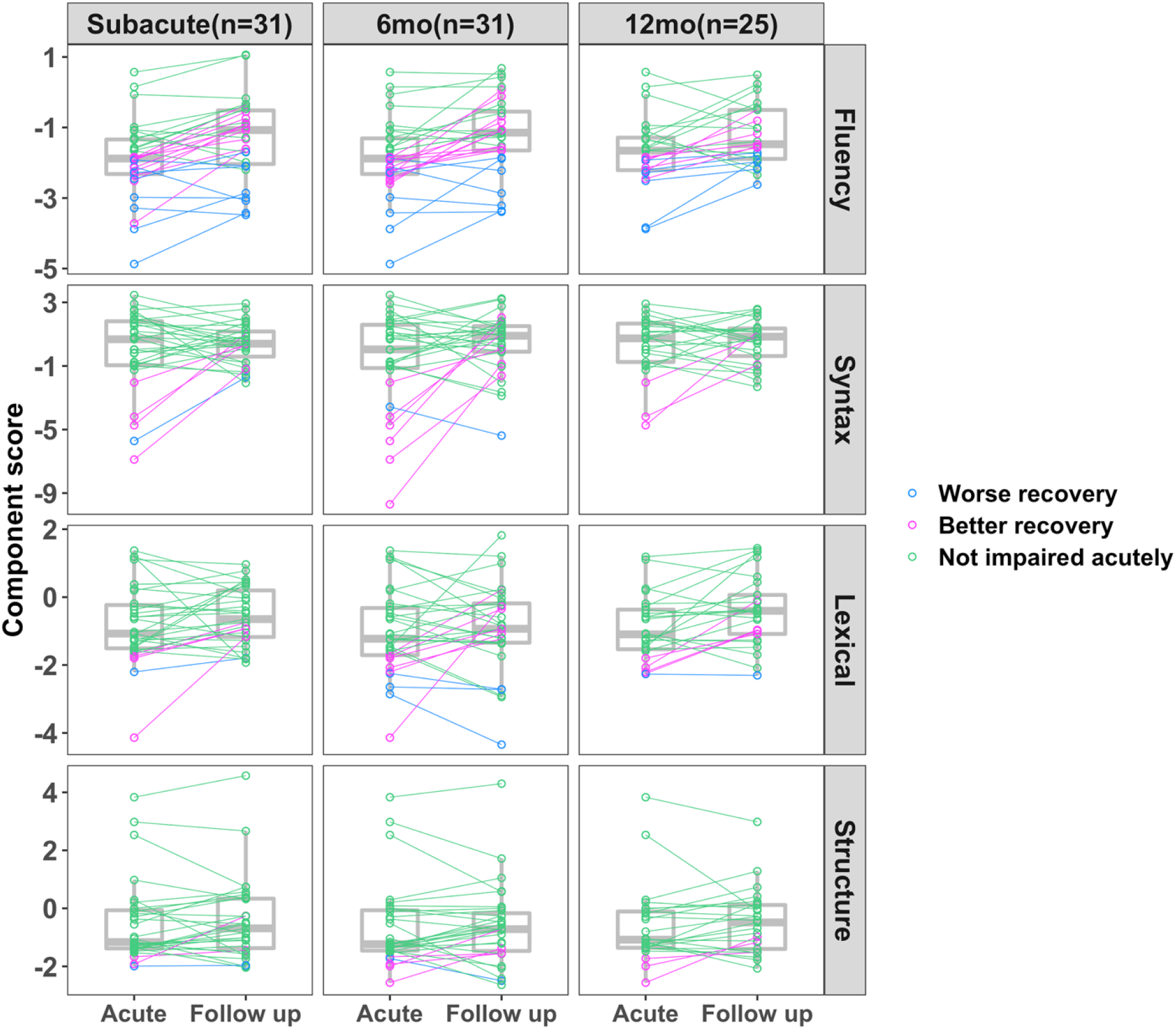
Recovery of four aspects of connected speech from acute to three follow-up time points in the year following stroke. Worse recovery was defined as both acute performance worse than 90% of controls’ performance (z’s < −1.67) and follow up performance still worse than 90% of controls (z’s < −1.67). In contrast, better recovery was defined as those with impaired acute performance z’s < −1.67 but subsequent follow up performance better than 90% of control performance (z’s > −1.67). We defined not impaired as individuals who performed better than 90% of controls’ performance acutely (z’s > −1.67).

**Figure 3.**
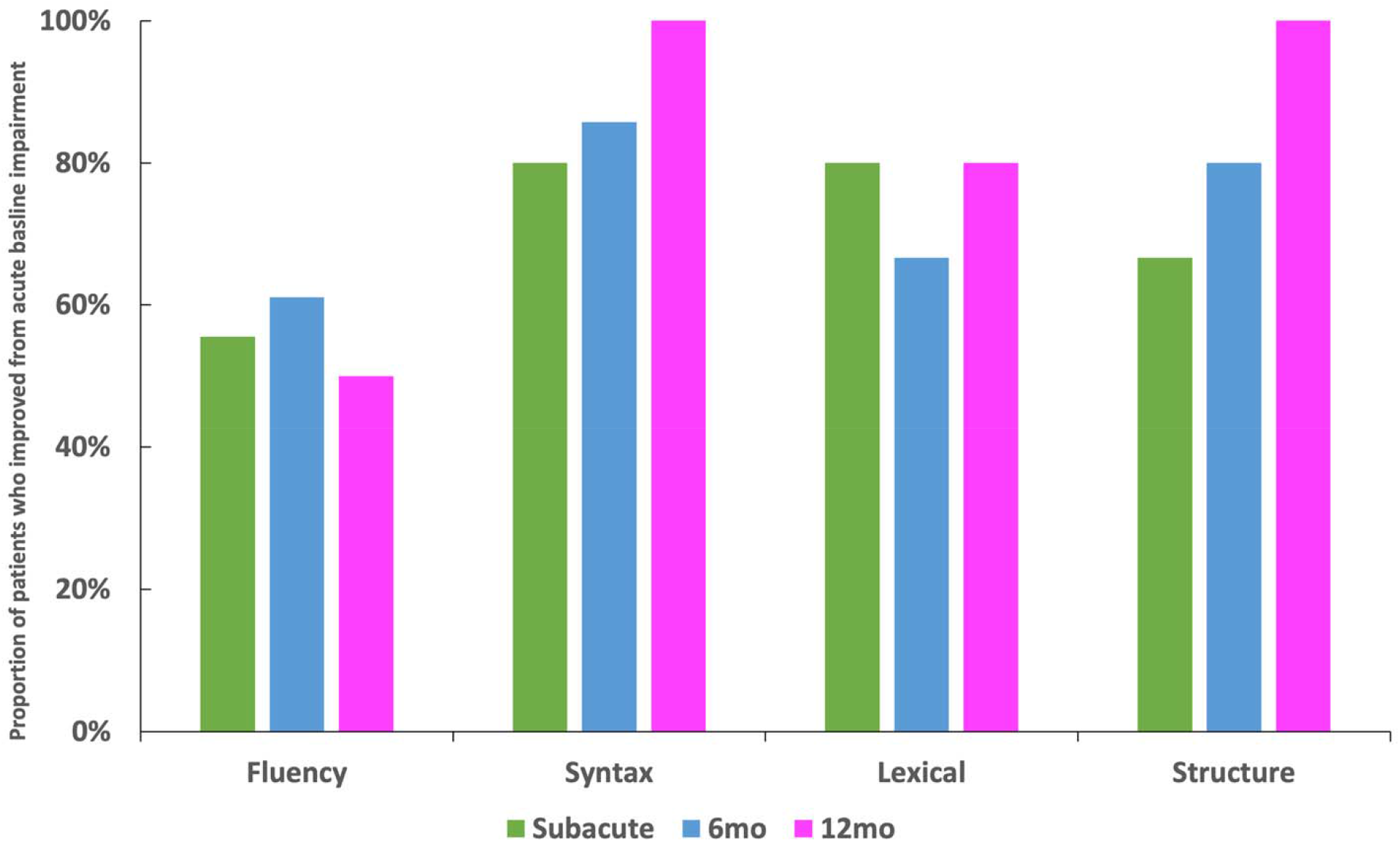
The proportions of patients who recovered to within normal levels (z > −1.67) after beginning with impairment acutely (z < −1.67) in four aspects of connected speech across three time points in the year following stroke.

Numbers outside parentheses denote the number of patients with impaired ability at the acute stage but within normal range at the follow-up stage (>-1.67) divided by the total number of patients with impaired abilities (<-1.67) at the acute stage. Numbers within parentheses reflect the proportion of patients who improved from acute impairment.

### Predicting connected speech recovery from acute white matter tract damage

Figure 4 displays the proportions of patients with acute damage to each white matter tract at each time point (see SI Table 2 for the summary of individual’s tract damage proportion). Among the seven tracts we examined, LSAF, FAT and IFOF were damaged in the most patients (n’s >= 8; 29-45% of patients), while the PSAF and UF had damage to the fewest patients (n’s =< 7; 16%-23% of patients). This pattern was also similar for those who were tested across all timepoints (LSAF, FAT, and IFOF n’s >= 6, 30%-35% of patients and for the PSAF and UF, n’s =< 4, 15%-20% of patients; see SI Figure 4).

**Figure 4.**
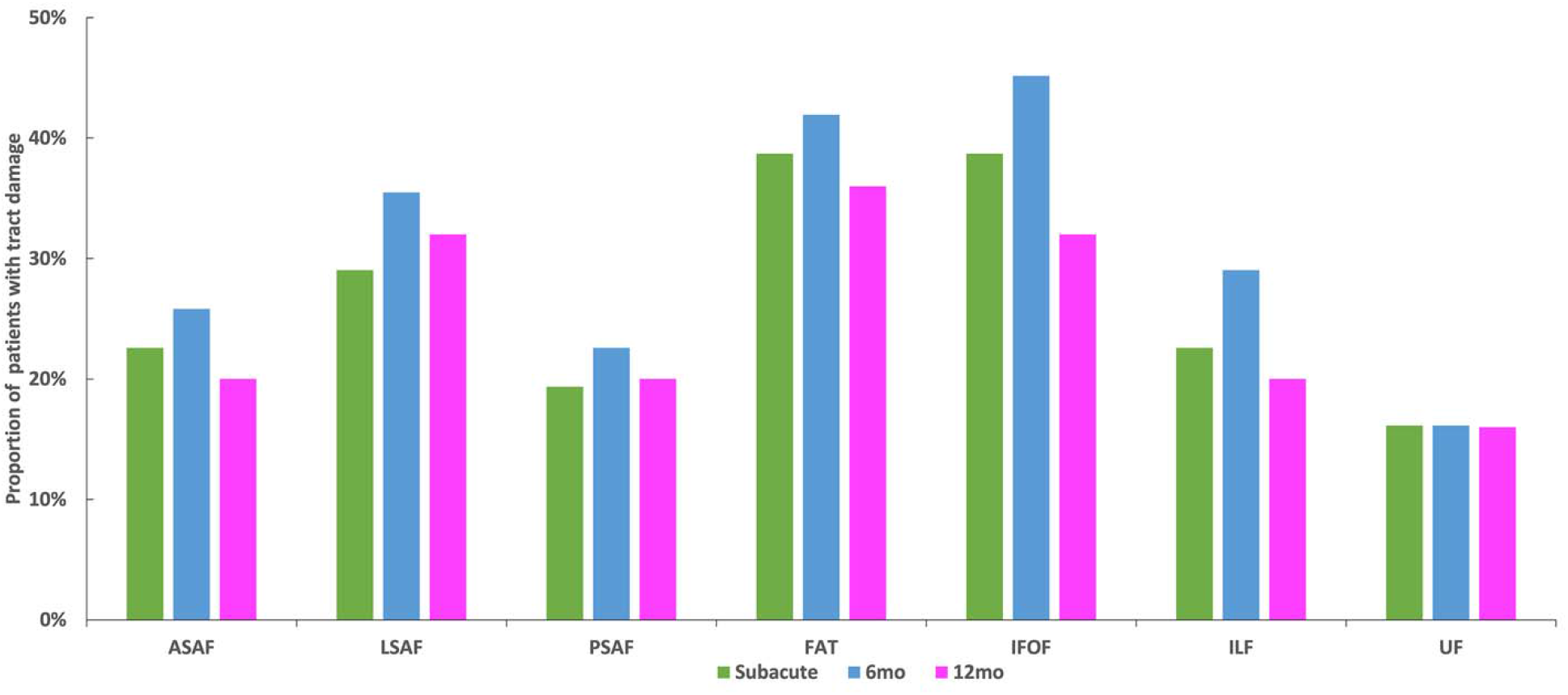
Proportion patients with tract damage for each tract across overlapping patient cohorts at subacute, early chronic (6mo) and late chronic (12mo) time points. ASAF/LSAF/PSAF: anterior/long/posterior segment of arcuate fasciculus; FAT: frontal aslant tract; IFOF: inferior fronto-occipital fasciculus; ILF: inferior longitudinal fasciculus; UF: uncinate fasciculus.

To identify factors potentially confounding relationships between tract damage and connected speech measurement recovery, we examined relationships between degree of recovery across time points and demographic variables (age, education, days tested post-stroke), total lesion volume and acute performance (see SI Table 3). The only significant variable associated with degree of connected speech recovery after multiple-comparisons correction (Bonferroni corrected *P* < 0.05/5) was acute baseline performance. Acute performance for all connected speech measures strongly correlated with recovery degree (r values < −0.32; *P* values < 0.08) except recovery of structural complexity at the subacute stage (r(29) = −0.24, *P* = 0.20) suggesting that patients with more severe impairments had more room for improvement. As a result, we included acute performance as a confounding variable in subsequent analyses. Acute lesion volume was not significantly related to either connected speech performance acutely, nor its recovery in either patient sample, except for acute fluency severity (r(29) = −0.36, *P* = 0.03).

Figure 5 and SI Table 4 show the lesion-symptom mapping results using white matter tract acute damage to predict connected speech recovery controlling for acute baseline performance and lesion volume. During the subacute stage after stroke, acute FAT damage led to decreased recovery of fluency at the subacute stage after stroke (z = 3.34, *P* = 0.001). In an overlapping subset of patients, acute FAT damage led to significantly decreased recovery of structural complexity at the chronic stages (6–12 months post-stroke; z values > 2.72, *P* values < 0.007). Results remained significant after controlling for the damage of FAT’s gray matter termini (subacute: *P* = 0.007; 6 months: *P* = 0.02; 12 months: *P* = 0.01). See SI Results and SI Figure 5 for uncorrected results. For 20 participants tested across all time points, effects of acute FAT damage on connected speech recovery at the chronic stages remained significant (z values > 2.14, *P* values < 0.03), while the effect at the subacute stage was not (z = 0.88, *P* = 0.19). See SI Table 5 for results. In sum, acute damage to the FAT contributed to decreased recovery of connected speech abilities at different times points after stroke.

**Figure 5.**
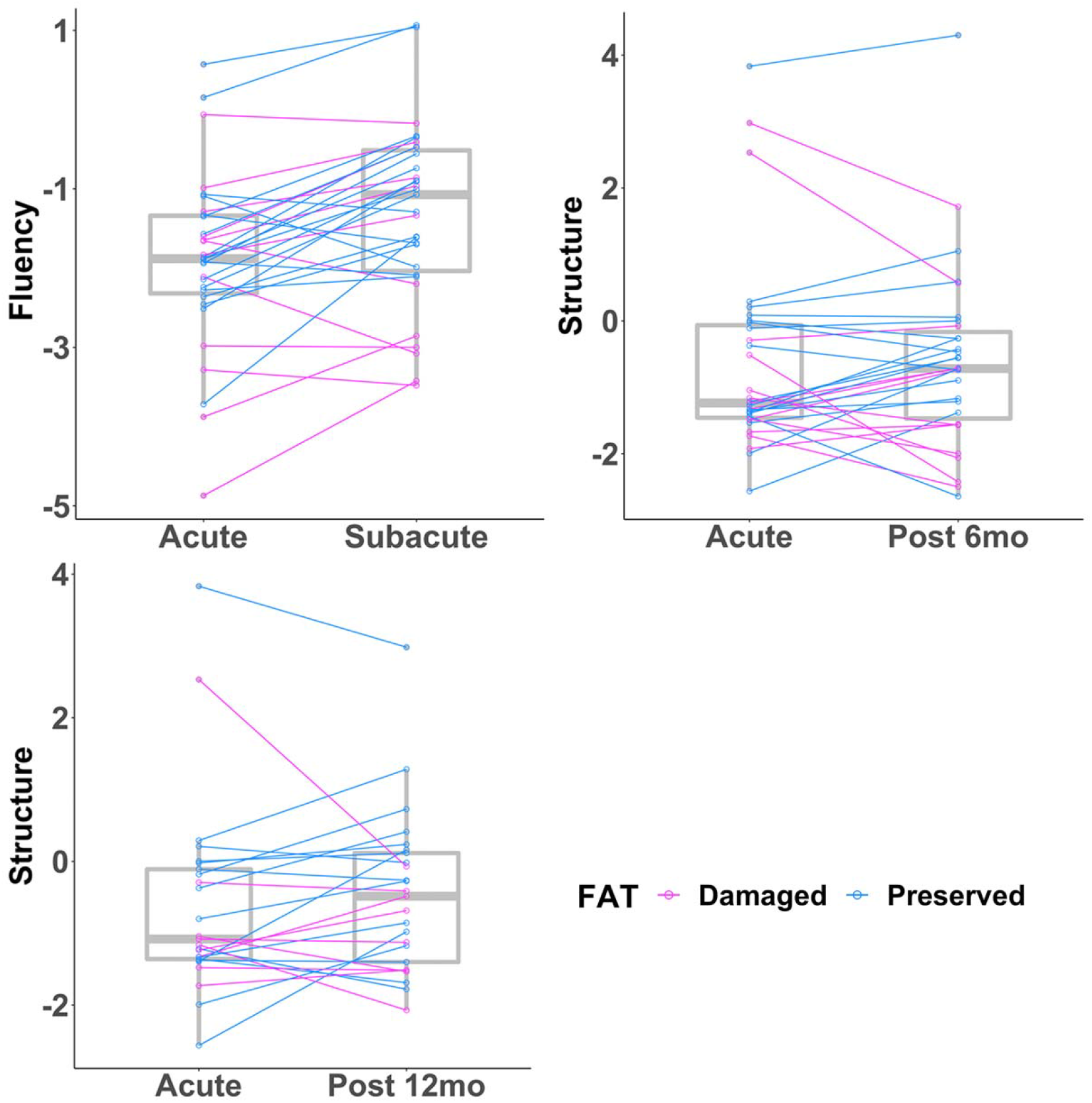
The white matter tracts whose acute damage predicted decreased recovery in four aspects of connected speech from the acute stage to three time points in the year following stroke after multiple-comparison correction. The box plots show the acute and follow-up connected speech scores clustered by tract damage. Magenta lines indicate subjects with damaged tracts, while blue lines indicate subjects with spared tracts.

## Discussion

We report the first large-scale longitudinal study of the impact of acute white matter damage on recovery of connected speech from the acute to chronic stages of left hemisphere stroke. The most frequent impairments in connected speech during the first year of recovery in those able to produce measurable connected speech acutely were reduced words per minute (∼25% of the cohort) followed by reduced ability to produce nouns in comparison to other types of words, decreased syntactic accuracy and reduced structural complexity (< ∼10%). White matter damage measured within the first week of stroke to a left dorsal pathway, the frontal aslant tract predicted worse subacute recovery of fluency and decreased chronic recovery of the structural complexity of connected speech. Importantly, acute white matter integrity predicted recovery of connected speech after controlling for overall lesion volume, damage to tract adjacent gray matter regions, and baseline acute connected speech performance. Our results provide novel evidence that recovery of connected speech after stroke depends on acute connectivity within frontal cortical regions.

### Connected speech impairments and recovery

Different aspects of connected speech production improved following the acute stage of stroke. To quantify connected speech, we used Ding et al.’s^5^ PCA coefficients generated for four connected speech components to calculate component scores for each participant. Of those who returned subacutely following stroke, reduced production of words per minute, i.e. fluency affected 58% of these subjects acutely. Approximately half of the impaired cohort recovered to within neurotypical fluency performance by the subacute stage of stroke. However, the remaining proportion of subjects with fluency impairments did not markedly change over the next year, suggesting a long-lasting deficit.

A second impairment in connected speech was reduced ability to produce nouns in comparison to other types of words. At the acute stage, 16% of all participants demonstrated lexical selection impairments. Of those, 80% recovered to neurotypical levels by the subacute stage. By a year post-stroke, this recovery proportion remained constant (80%).

Recovery was better for other aspects of connected speech. The ability to produce syntactically accurate speech, including more well-formed sentences, more words within as opposed to outside of sentences and increased production of required determiners was impaired acutely in 12% of all participants but by the late chronic stage all subjects who demonstrated syntactic impairment recovered to within a neurotypical range of performance. Similarly of 12% who suffered acute impairment producing structurally complex speech including elaborated phrases, sentence embeddings, and longer sentences, all participants impaired acutely recovered by the late chronic stage.

Overall, of participants with acute connected speech impairments, connected speech improved in over 50% of returning participants by the subacute stage of stroke. Encouragingly, nearly all participants impaired acutely improved their connected speech by the late chronic stage of stroke, except for fluency, which remained impaired in almost half the cohort. This pattern was similar in those participants who were tested across all the timepoints. It is important to note that we enrolled patients who could produce measurable connected speech independent of any clinical diagnosis of language impairment. Thus, it is striking that even when not including patients with severe production deficits, only half of those with impaired fluency recovered. In comparison, more than 80% of participants impaired acutely produced syntactically accurate, structurally complex, and lexically diverse connected speech by the late chronic stage. However, it remains an open question the degree to which more severe connected speech deficits follow the connected speech patterns of improvement observed here.

### Acute white matter tract damage and connected speech recovery

We examined how damage to white matter tracts at the acute phase of stroke impacted the recovery of connected speech during reorganization of function across the subacute, early chronic and late chronic phases of stroke. Of several dorsal and ventral white matter tracts we examined across different stages of recovery, the FAT was the only tract significantly related with connected speech recovery after multiple comparison correction. At the subacute stage of stroke, acute FAT damage decreased degree of fluency recovery as expected. However, in the subset of participants who were able to return across all time points we did not replicate this association. Also, we did not find that acute damage to tracts connecting frontal with posterior regions including the anterior segment of the arcuate fasciculus or a ventral tract, the uncinate fasciculus predicted fluency impairments as has been demonstrated in chronic aphasia.^22,59^ That acute FAT damage reduced early recovery of fluency during connected speech is consistent with evidence in chronic aphasia demonstrating that FAT integrity is critical for fluency as measured by words per minute during connected speech (albeit not performing direct statistical analysis;^3^ cf.^60^ for similar results in primary progressive aphasia), qualitative assessment of fluency^59^ and word generation during categorical fluency tasks.^61,62^ Conflicting relationships between anterior white matter damage and fluency performance may be in part because of both how fluency is measured as well as the multidimensionality of fluency. Previous studies’ positive results measured fluency with a qualitative 10-pt scale assessment (via the Western Aphasia Battery^63^) which captures other functions including prosody, syntax, and lexical retrieval.^59^ Additionally complicating the picture, fluency is dependent on multiple capacities (e.g., articulation, phonology, comprehension, working memory; cf.^64^). Thus, after stroke, a speaker will have reduced fluency because of a deficit in any or all these capacities. By removing participants unable to return across all time points, we may have removed a significant proportion of participants with a common underlying cause of disfluency associated with acute FAT damage. Understanding the behavioral and neuroanatomical contributions to disfluency after stroke will clearly continue to be important directions to address in future research.

The second result we observed was at the early and late chronic stages of stroke, where acute FAT damage also decreased degree of structural complexity recovery. This pattern replicated in participants who returned across all timepoints. These results were inconsistent with our predictions as only damage to left temporal-parietal regions distant from the FAT has been associated with impairments in the structural complexity of speech, both acutely^5^ and chronically.^10,65–67^ Thus, this latter result suggests that cortical regions associated with the frontal aslant tract, like the IFG, may functionally reorganize to support posterior temporo-parietal processes involved in creating more structurally complex connected speech in chronic stage. Together, these results demonstrate the critical role of frontal connectivity for connected speech during reorganization of function.

### Other predictors of connected speech recovery

Lastly, our analysis found that acute lesion volume showed a relationship with the deficit severity of acute stage connected speech fluency, but with no other components of connected speech. Moreover, it is important to note that lesion volume did not predict the degree of connected speech recovery at any time point in the year after stroke. Although lesion volume often predicts language deficit severity in stroke (e.g.^6,19,34,36,68–70^; cf.^18^), lesion volume does not predict the degree of general language^71^ or domain-specific cognitive function recovery.^72^ Instead, initial performance was more important to the recovery degree (cf.^71,72^). We also found acute performance a significant diagnostic of connected speech recovery outcomes which is consistent with longitudinal recovery patterns of upper-extremity movement after stroke.^73^

## Limitations

There are several inherent limitations which arise as a result of the difficulty in conducting large-scale longitudinal analyses of recovery from stroke. First, even though participants significantly overlapped across the subacute, early and late chronic timepoints (between 75-84% of participants), for a variety of reasons not every patient was tested at all follow-up time points. For example, the post 12 months group included fewer individuals and individuals with damage to white matter tracts. We speculate participants with higher tract damage are more likely not to attend further follow-up studies, probably because they suffer more difficulties due to the severe damage. However, to maximize power to detect factors which affected recovery, we still included the largest number of subjects tested both acutely and at a single subsequent time point. Although this approach helped avoid type II statistical errors, i.e. false negatives, it rendered direct comparisons between time points difficult to interpret. We addressed this limitation by examining the subset of participants who were able to return across all time points. These analyses confirmed that the FAT is critical to the recovery of connected speech. Second, in our cohort, there was a low prevalence of participants with severe impairments in the syntactic accuracy or structural complexity of their connected speech. We conjecture that patients with these types of severe connected speech impairments could not produce measurable spontaneous speech acutely, an exclusion criteria for this study. To understand whether different recovery mechanisms occur in those with more severe connected speech deficits, our laboratory is engaged in an ongoing effort to expand subject recruitment to these clinical populations. Finally, future work will test the degree to which the right hemisphere contributes to recovery of connected speech after stroke as evidence suggests its role in language recovery (cf.^20,69,74^).

## Conclusion

This longitudinal analysis from acute to chronic stroke provides the first evidence that recovery of fluent and structurally complex spontaneous connected speech requires intact left frontal connectivity via the frontal aslant tract. That the frontal aslant tract was critical for recovery at early as well as later stages of stroke demonstrates it plays a lasting and important role for the reorganization of function related to the successful production of connected speech. These results suggest that measures of acute cortical disconnection may be useful biomarkers to identify patients who will most benefit from early interventions to remediate chronic connected speech impairments.

## Data Availability

All data produced in the present study are available upon reasonable request to the authors.

## Abbreviations

ASAF/LSAF/PSAF: anterior/long/posterior segment of arcuate fasciculus;
FAT: frontal aslant tract;
IFG: inferior frontal gyrus;
IFOF: inferior fronto-occipital fasciculus;
ILF: inferior longitudinal fasciculus;
PCA: principal component analysis;
QPA: quantitative production analysis;
UF: uncinate fasciculus

## Acknowledgements

The authors wish to thank Jolie Anderson, Miranda Brenneman, Cris Hamilton, Danielle Rossi, and Chia-Ming Lei for data collection. We wish to thank Erica Johns, Bowie Lin, Hao Yan, Rachel Zahn, and Riya Mehta for transcription and analysis of narrative speech samples. We thank the clinical neurological intensive care unit teams at the University of Texas Health Sciences Center and Memorial Hermann Hospital, The Houston Methodist Hospital, and the Baylor St. Luke’s Hospital for their assistance in patient recruitment and neurological assessment. We gratefully acknowledge and thank our research subjects and their caregivers for their willingness to participate in this research. This work was presented at the Society for the Neurobiology of Language (2021).

## References

1. Mirman D, Chen Q, Zhang Y, et al. Neural organization of spoken language revealed by lesion–symptom mapping. Nat Commun. 2015;6(1):6762. doi:10.1038/ncomms7762

2. Schwartz MF, Dell GS, Martin N, Gahl S, Sobel P. A case-series test of the interactive two-step model of lexical access: Evidence from picture naming. J Mem Lang. 2006;54(2):228–264. doi:10.1016/j.jml.2005.10.001

3. Alyahya RSW, Halai AD, Conroy P, Lambon Ralph MA. A unified model of post-stroke language deficits including discourse production and their neural correlates. Brain. 2020;143(5):1541–1554. doi:10.1093/brain/awaa074

4. Borovsky A, Saygin AP, Bates E, Dronkers N. Lesion correlates of conversational speech production deficits. Neuropsychologia. 2007;45(11):2525–2533. doi:10.1016/j.neuropsychologia.2007.03.023

5. Ding J, Martin RC, Hamilton AC, Schnur TT. Dissociation between frontal and temporal-parietal contributions to connected speech in acute stroke. Brain. 2020;143(3):862–876. doi:10.1093/brain/awaa027

6. Halai AD, Woollams AM, Lambon Ralph MA. Using principal component analysis to capture individual differences within a unified neuropsychological model of chronic post-stroke aphasia: Revealing the unique neural correlates of speech fluency, phonology and semantics. Cortex. 2017;86:275–289. doi:10.1016/j.cortex.2016.04.016

7. Hillis AE, Beh YY, Sebastian R, et al. Predicting recovery in acute poststroke aphasia. Ann Neurol. 2018;83(3):612–622. doi:10.1002/ana.25184

8. Meier EL, Johnson JP, Pan Y, Kiran S. The utility of lesion classification in predicting language and treatment outcomes in chronic stroke-induced aphasia. Brain Imaging Behav. 2019;13(6):1510–1525. doi:10.1007/s11682-019-00118-3

9. van Hees S, McMahon K, Angwin A, de Zubicaray G, Read S, Copland DA. Changes in White Matter Connectivity Following Therapy for Anomia Post stroke. Neurorehabil Neural Repair. 2014;28(4):325–334. doi:10.1177/1545968313508654

10. Matchin W, Basilakos A, Stark BC, den Ouden DB, Fridriksson J, Hickok G. Agrammatism and Paragrammatism: A Cortical Double Dissociation Revealed by Lesion-Symptom Mapping. Neurobiol Lang. 2020;1(2):208–225. doi:10.1162/nol_a_00010

11. Gajardo-Vidal A, Lorca-Puls DL, team P, et al. Damage to Broca’s area does not contribute to long-term speech production outcome after stroke. Brain. 2021;144(3):817–832. doi:10.1093/brain/awaa460

12. Stark BC, Basilakos A, Hickok G, Rorden C, Bonilha L, Fridriksson J. Neural organization of speech production: A lesion-based study of error patterns in connected speech. Cortex. 2019;117:228–246. doi:10.1016/j.cortex.2019.02.029

13. Siegel JS, Ramsey LE, Snyder AZ, et al. Disruptions of network connectivity predict impairment in multiple behavioral domains after stroke. Proc Natl Acad Sci. 2016;113(30):E4367–E4376. doi:10.1073/pnas.1521083113

14. Thiebaut de Schotten M, Foulon C, Nachev P. Brain disconnections link structural connectivity with function and behaviour. Nat Commun. 2020;11(1):5094. doi:10.1038/s41467-020-18920-9

15. Reber J, Hwang K, Bowren M, et al. Cognitive impairment after focal brain lesions is better predicted by damage to structural than functional network hubs. Proc Natl Acad Sci. 2021;118(19). doi:10.1073/pnas.2018784118

16. Catani M, Jones DK, Ffytche DH. Perisylvian language networks of the human brain. Ann Neurol. 2005;57(1):8–16. doi:10.1002/ana.20319

17. Bernard F, Zemmoura I, Ter Minassian A, Lemée JM, Menei P. Anatomical variability of the arcuate fasciculus: a systematical review. Surg Radiol Anat. 2019;41(8):889–900. doi:10.1007/s00276-019-02244-5

18. Marchina S, Zhu LL, Norton A, Zipse L, Wan CY, Schlaug G. Impairment of Speech Production Predicted by Lesion Load of the Left Arcuate Fasciculus. Stroke. 2011;42(8):2251–2256. doi:10.1161/STROKEAHA.110.606103

19. Wang J, Marchina S, Norton A, Wan C, Schlaug G. Predicting speech fluency and naming abilities in aphasic patients. Front Hum Neurosci. 2013;7. Accessed January 19, 2022. https://www.frontiersin.org/article/10.3389/fnhum.2013.00831

20. Pani E, Zheng X, Wang J, Norton A, Schlaug G. Right hemisphere structures predict poststroke speech fluency. Neurology. 2016;86(17):1574–1581. doi:10.1212/WNL.0000000000002613

21. Keser Z, Meier EL, Stockbridge MD, Hillis AE. The role of microstructural integrity of major language pathways in narrative speech in the first year after stroke. J Stroke Cerebrovasc Dis. 2020;29(9):105078. doi:10.1016/j.jstrokecerebrovasdis.2020.105078

22. Fridriksson J, Guo D, Fillmore P, Holland A, Rorden C. Damage to the anterior arcuate fasciculus predicts non-fluent speech production in aphasia. Brain. 2013;136(11):3451–3460. doi:10.1093/brain/awt267

23. Von Der Heide RJ, Skipper LM, Klobusicky E, Olson IR. Dissecting the uncinate fasciculus: disorders, controversies and a hypothesis. Brain. 2013;136(6):1692–1707. doi:10.1093/brain/awt094

24. Catani M, Dell’Acqua F, Vergani F, et al. Short frontal lobe connections of the human brain. Cortex. 2012;48(2):273–291. doi:10.1016/j.cortex.2011.12.001

25. Rojkova K, Volle E, Urbanski M, Humbert F, Dell’Acqua F, Thiebaut de Schotten M. Atlasing the frontal lobe connections and their variability due to age and education: a spherical deconvolution tractography study. Brain Struct Funct. 2016;221(3):1751–1766. doi:10.1007/s00429-015-1001-3

26. Martino J, Brogna C, Robles SG, Vergani F, Duffau H. Anatomic dissection of the inferior fronto-occipital fasciculus revisited in the lights of brain stimulation data. Cortex. 2010;46(5):691–699. doi:10.1016/j.cortex.2009.07.015

27. Wu Y, Sun D, Wang Y, Wang Y. Subcomponents and Connectivity of the Inferior Fronto-Occipital Fasciculus Revealed by Diffusion Spectrum Imaging Fiber Tracking. Front Neuroanat. 2016;10. Accessed January 19, 2022. https://www.frontiersin.org/article/10.3389/fnana.2016.00088

28. Stockbridge MD, Berube S, Goldberg E, et al. Differences in linguistic cohesion within the first year following right-and left-hemisphere lesions. Aphasiology. 2021;35(3):357–371. doi:10.1080/02687038.2019.1693026

29. Hoffman P, Cogdell-Brooke L, Thompson HE. Going off the rails: Impaired coherence in the speech of patients with semantic control deficits. Neuropsychologia. 2020;146:107516. doi:10.1016/j.neuropsychologia.2020.107516

30. Rochon E, Saffran EM, Berndt RS, Schwartz MF. Quantitative Analysis of Aphasic Sentence Production: Further Development and New Data. Brain Lang. 2000;72(3):193–218. doi:10.1006/brln.1999.2285

31. Fromm D, Katta S, Paccione M, et al. A Comparison of Manual Versus Automated Quantitative Production Analysis of Connected Speech. J Speech Lang Hear Res. 2021;64(4):1271–1282. doi:10.1044/2020_JSLHR-20-00561

32. Ochfeld E, Newhart M, Molitoris J, et al. Ischemia in Broca Area Is Associated With Broca Aphasia More Reliably in Acute Than in Chronic Stroke. Stroke. 2010;41(2):325–330. doi:10.1161/STROKEAHA.109.570374

33. Shahid H, Sebastian R, Schnur TT, et al. Important considerations in lesion-symptom mapping: Illustrations from studies of word comprehension. Hum Brain Mapp. 2017;38(6):2990–3000. doi:10.1002/hbm.23567

34. Yourganov G, Fridriksson J, Rorden C, Gleichgerrcht E, Bonilha L. Multivariate Connectome-Based Symptom Mapping in Post-Stroke Patients: Networks Supporting Language and Speech. J Neurosci. 2016;36(25):6668–6679. doi:10.1523/JNEUROSCI.4396-15.2016

35. Saur D, Lange R, Baumgaertner A, et al. Dynamics of language reorganization after stroke. Brain. 2006;129(6):1371–1384. doi:10.1093/brain/awl090

36. Stockert A, Wawrzyniak M, Klingbeil J, et al. Dynamics of language reorganization after left temporo-parietal and frontal stroke. Brain. 2020;143(3):844–861. doi:10.1093/brain/awaa023

37. Stefaniak JD, Halai AD, Lambon Ralph MA. The neural and neurocomputational bases of recovery from post-stroke aphasia. Nat Rev Neurol. 2020;16(1):43–55. doi:10.1038/s41582-019-0282-1

38. Lazar RM, Minzer B, Antoniello D, Festa JR, Krakauer JW, Marshall RS. Improvement in Aphasia Scores After Stroke Is Well Predicted by Initial Severity. Stroke. 2010;41(7):1485–1488. doi:10.1161/STROKEAHA.109.577338

39. Saur D, Ronneberger O, Kümmerer D, Mader I, Weiller C, Klöppel S. Early functional magnetic resonance imaging activations predict language outcome after stroke. Brain. 2010;133(4):1252–1264. doi:10.1093/brain/awq021

40. Martin RC, Schnur TT. Independent contributions of semantic and phonological working memory to spontaneous speech in acute stroke. Cortex. 2019;112:58–68. doi:10.1016/j.cortex.2018.11.017

41. Martin RC, Ding J, Hamilton AC, Schnur TT. Working Memory Capacities Neurally Dissociate: Evidence from Acute Stroke. Cereb Cortex Commun. 2021;2(2):tgab005. doi:10.1093/texcom/tgab005

42. Corbetta M, Ramsey L, Callejas A, et al. Common Behavioral Clusters and Subcortical Anatomy in Stroke. Neuron. 2015;85(5):927–941. doi:10.1016/j.neuron.2015.02.027

43. Meier EL, Sheppard SM, Goldberg EB, et al. Naming errors and dysfunctional tissue metrics predict language recovery after acute left hemisphere stroke. Neuropsychologia. 2020;148:107651. doi:10.1016/j.neuropsychologia.2020.107651

44. Folstein MF, Robins LN, Helzer JE. The Mini-Mental State Examination. Arch Gen Psychiatry. 1983;40(7):812. doi:10.1001/archpsyc.1983.01790060110016

45. Ehrlich A, Perrault C, Jeffers S. Cinderella. Dutton Children’s Books Children’s; 2004.

46. Gordon JK. A quantitative production analysis of picture description. Aphasiology. 2006;20(2-4):188–204. doi:10.1080/02687030500472777

47. Saffran EM, Berndt RS, Schwartz MF. The quantitative analysis of agrammatic production: Procedure and data. Brain Lang. 1989;37(3):440–479. doi:10.1016/0093-934X(89)90030-8

48. Cox RW. AFNI: Software for Analysis and Visualization of Functional Magnetic Resonance Neuroimages. Comput Biomed Res. 1996;29(3):162–173. doi:10.1006/cbmr.1996.0014

49. Yushkevich PA, Piven J, Hazlett HC, et al. User-guided 3D active contour segmentation of anatomical structures: Significantly improved efficiency and reliability. NeuroImage. 2006;31(3):1116–1128. doi:10.1016/j.neuroimage.2006.01.015

50. Avants BB, Epstein CL, Grossman M, Gee JC. Symmetric diffeomorphic image registration with cross-correlation: Evaluating automated labeling of elderly and neurodegenerative brain. Med Image Anal. 2008;12(1):26–41. doi:10.1016/j.media.2007.06.004

51. Catani M, Allin MPG, Husain M, et al. Symmetries in human brain language pathways correlate with verbal recall. Proc Natl Acad Sci. 2007;104(43):17163–17168. doi:10.1073/pnas.0702116104

52. Forkel SJ, Rogalski E, Sancho ND, et al. Anatomical evidence of an indirect pathway for word repetition. Neurology. 2020;94(6):e594–e606. doi:10.1212/WNL.0000000000008746

53. López-Barroso D, Catani M, Ripollés P, Dell’Acqua F, Rodríguez-Fornells A, Diego-Balaguer R de. Word learning is mediated by the left arcuate fasciculus. Proc Natl Acad Sci. 2013;110(32):13168–13173. doi:10.1073/pnas.1301696110

54. DeMarco AT, Turkeltaub PE. A multivariate lesion symptom mapping toolbox and examination of lesion-volume biases and correction methods in lesion-symptom mapping. Hum Brain Mapp. 2018;39(11):4169–4182. doi:10.1002/hbm.24289

55. Sperber C, Karnath HO. On the validity of lesion-behaviour mapping methods. Neuropsychologia. 2018;115:17–24. doi:10.1016/j.neuropsychologia.2017.07.035

56. Pustina D, Avants B, Faseyitan OK, Medaglia JD, Coslett HB. Improved accuracy of lesion to symptom mapping with multivariate sparse canonical correlations. Neuropsychologia. 2018;115:154–166. doi:10.1016/j.neuropsychologia.2017.08.027

57. Lacey EH, Skipper-Kallal LM, Xing S, Fama ME, Turkeltaub PE. Mapping Common Aphasia Assessments to Underlying Cognitive Processes and Their Neural Substrates. Neurorehabil Neural Repair. 2017;31(5):442–450. doi:10.1177/1545968316688797

58. Tzourio-Mazoyer N, Landeau B, Papathanassiou D, et al. Automated Anatomical Labeling of Activations in SPM Using a Macroscopic Anatomical Parcellation of the MNI MRI Single-Subject Brain. NeuroImage. 2002;15(1):273–289. doi:10.1006/nimg.2001.0978

59. Basilakos A, Fillmore PT, Rorden C, Guo D, Bonilha L, Fridriksson J. Regional White Matter Damage Predicts Speech Fluency in Chronic Post-Stroke Aphasia. Front Hum Neurosci. 2014;8. Accessed January 19, 2022. https://www.frontiersin.org/article/10.3389/fnhum.2014.00845

60. Catani M, Mesulam MM, Jakobsen E, et al. A novel frontal pathway underlies verbal fluency in primary progressive aphasia. Brain. 2013;136(8):2619–2628. doi:10.1093/brain/awt163

61. Li M, Zhang Y, Song L, et al. Structural connectivity subserving verbal fluency revealed by lesion-behavior mapping in stroke patients. Neuropsychologia. 2017;101:85–96. doi:10.1016/j.neuropsychologia.2017.05.008

62. Foulon C, Cerliani L, Kinkingnéhun S, et al. Advanced lesion symptom mapping analyses and implementation as BCBtoolkit. GigaScience. 2018;7(3):giy004. doi:10.1093/gigascience/giy004

63. Kertesz A. Western Aphasia Battery--Revised. Published online 2006. doi:10.1037/t15168-000

64. Nozari N, Faroqi-Shah Y. Investigating the origin of nonfluency in aphasia: A path modeling approach to neuropsychology. Cortex. 2017;95:119–135. doi:10.1016/j.cortex.2017.08.003

65. den Ouden DB, Malyutina S, Basilakos A, et al. Cortical and structural-connectivity damage correlated with impaired syntactic processing in aphasia. Hum Brain Mapp. 2019;40(7):2153–2173. doi:10.1002/hbm.24514

66. Lukic S, Bonakdarpour B, Ouden DD, Price C, Thompson C. Neural Mechanisms of Verb and Sentence Production: A Lesion-deficit Study. Procedia -Soc Behav Sci. 2013;94:34–35. doi:10.1016/j.sbspro.2013.09.014

67. Henseler I, Regenbrecht F, Obrig H. Lesion correlates of patholinguistic profiles in chronic aphasia: comparisons of syndrome-, modality-and symptom-level assessment. Brain. 2014;137(3):918–930. doi:10.1093/brain/awt374

68. Ivanova MV, Zhong A, Turken A, Baldo JV, Dronkers NF. Functional Contributions of the Arcuate Fasciculus to Language Processing. Front Hum Neurosci. 2021;15. Accessed January 19, 2022. https://www.frontiersin.org/article/10.3389/fnhum.2021.672665

69. Forkel SJ, Thiebaut de Schotten M, Dell’Acqua F, et al. Anatomical predictors of aphasia recovery: a tractography study of bilateral perisylvian language networks. Brain. 2014;137(7):2027–2039. doi:10.1093/brain/awu113

70. Hope TMH, Seghier ML, Leff AP, Price CJ. Predicting outcome and recovery after stroke with lesions extracted from MRI images. NeuroImage Clin. 2013;2:424–433. doi:10.1016/j.nicl.2013.03.005

71. Laska AC, Hellblom A, Murray V, Kahan T, Von Arbin M. Aphasia in acute stroke and relation to outcome. J Intern Med. 2001;249(5):413–422. doi:10.1046/j.1365-2796.2001.00812.x

72. Nys GMS, Zandvoort MJEV, Kort Plmd, et al. Domain-specific cognitive recovery after first-ever stroke: A follow-up study of 111 cases. J Int Neuropsychol Soc. 2005;11(7):795–806. doi:10.1017/S1355617705050952

73. van der Vliet R, Selles RW, Andrinopoulou ER, et al. Predicting Upper Limb Motor Impairment Recovery after Stroke: A Mixture Model. Ann Neurol. 2020;87(3):383–393. doi:10.1002/ana.25679

74. Schlaug G, Marchina S, Norton A. Evidence for Plasticity in White-Matter Tracts of Patients with Chronic Broca’s Aphasia Undergoing Intense Intonation-based Speech Therapy. Ann N Y Acad Sci. 2009;1169(1):385–394. doi:10.1111/j.1749-6632.2009.04587.x

