## Supplementary information for "Anterior Connectivity Critical for Recovery of Connected Speech after Stroke"

**SI Results**

**Lesion-symptom mapping results significant at the *P* < .05 threshold.** Several Lesion-symptom mapping findings were significant but did not survive correction for multiple comparisons (see SI Table 4). At the subacute stage, acute FAT damage predicted decreased recovery of syntactic abilities (z = 1.91, *P* = 0.03). Chronically, decreased recovery of syntax 6 months (z =2.91, *P* = 0.01) and 12 months post stroke (z = 2.73, *P* = 0.01) was associated with acute damage to posterior and long segments of the AF. We replicated these findings when additionally controlling for the degree of gray matter damage at the origins and termini for each tract (FAT: *P* = 0.03; LSAF: *P* = 0.02; save for the marginally significant relationship between acute PSAF damage and degree of syntactic complexity recovery at 6 months post-stroke, *P* = .08).

**SI Table 1. The number and proportion of patients testing at all the time points (n=20) who recovered to within normal levels (z > -1.67) after beginning with impairment acutely (z < -1.67) in four aspects of connected speech.**

|  | Fluency | Syntactic accuracy | Lexical selection | Structural  complexity |
| --- | --- | --- | --- | --- |
| Subacute (n = 20) | 5/9 (56%) | 3/3 (100%) | 1/2 (50%) | 1/1 (100%) |
| 6mo (n = 20) | 7/9 (78%) | 3/3 (100%) | 2/2 (100%) | 1/1 (100%) |
| 12mo (n = 20) | 5/9 (56%) | 3/3 (100%) | 2/2 (100%) | 1/1 (100%) |

Numbers outside parentheses denote the number of patients with impaired ability at the acute stage but within normal range at the follow-up stage (>-1.67) divided by the total number of patients with impaired abilities (<-1.67) at the acute stage. Numbers within parentheses reflect the proportion of patients who improved from acute impairment.

**SI Table 2.** **Summary of average acute damage proportion for each tract across patient cohorts at subacute, early chronic (6mo), and late chronic (12mo) time points.**

| Tract | Subacute (n = 31) | 6mo (n = 31) | 12mo (n = 25) | All time points (n=20) |
| --- | --- | --- | --- | --- |
| ASAF | 2%±4% (0-16%) | 2%±4% (0-16%) | 1%±2% (0-8%) | 1%±2% (0-8%) |
| LSAF | 2%±5% (0-23%) | 3%±6% (0-23%) | 1%±3% (0-10%) | 1%±3% (0-10%) |
| PSAF | 2%±9% (0-50%) | 4%±12% (0-50%) | 2%±6% (0-30%) | 1%±1% (0-5%) |
| FAT | 2%±5% (0-24%) | 2%±5% (0-24%) | 2%±5% (0-24%) | 2%±6% (0-24%) |
| IFOF | 1%±3% (0-11%) | 1%±2% (0-11%) | 1%±2% (0-11%) | 1%±3% (0-11%) |
| ILF | 1%±4% (0-16%) | 1%±4% (0-16%) | 1%±3% (0-16%) | 1%±4% (0-16%) |
| UF | 1%±2% (0-14%) | 1%±2% (0-14%) | 1%±3% (0-14%) | 1%±3% (0-14%) |

Numbers denotes Mean±S.D. (range).

**SI Table 3. Correlations between demographic, behavioral, lesion volume, and days tested after stroke variables and recovery in four aspects of connected speech from acute to three time points in the year following stroke.**

|  |  | Acute performance | Age | Education | Lesion Volume | Days after stroke onset |
| --- | --- | --- | --- | --- | --- | --- |
| Acute  (n = 38) | Fluency |  | -0.08 (0.64) | 0.28 (0.09) | -0.36 (0.03) ^#^ | -0.08 (0.65) |
|  | Syntactic accuracy |  | -0.32 (0.05) | 0.08 (0.64) | -0.22 (0.19) | 0.11 (0.53) |
|  | Lexical selection |  | -0.03 (0.85) | 0.19 (0.27) | -0.27 (0.10) | -0.16 (0.35) |
|  | Structural complexity |  | 0.18 (0.27) | 0.31 (0.07) | 0.06 (0.74) | -0.12 (0.49) |
| Subacute (n = 31) | Fluency | -0.32 (0.08) | -0.11 (0.56) | 0.00 (0.98) | 0.30 (0.10) | 0.24 (0.20) |
|  | Syntactic accuracy | -0.87 (2E-10)* | 0.24 (0.19) | -0.12 (0.53) | 0.22 (0.25) | 0.35 (0.05) |
|  | Lexical selection | -0.71 (7E-6)* | 0.14 (0.46) | -0.19 (0.31) | 0.21 (0.26) | -0.09 (0.61) |
|  | Structural complexity | -0.24 (0.20) | -0.14 (0.45) | 0.01 (0.94) | -0.26 (0.15) | -0.05 (0.79) |
| 6mo (n = 31) | Fluency | -0.43 (0.02)^#^ | 0.08 (0.68) | 0.31 (0.11) | 0.18 (0.34) | 0.27 (0.14) |
|  | Syntactic accuracy | -0.82 (2E-8)* | 0.10 (0.59) | -0.25 (0.19) | 0.23 (0.22) | 0.29 (0.12) |
|  | Lexical selection | -0.50 (0.004)* | 0.07 (0.69) | 0.004 (0.99) | 0.34 (0.06) | 0.03 (0.86) |
|  | Structural complexity | -0.39 (0.03)^#^ | 0.16 (0.40) | 0.17 (0.37) | -0.23 (0.21) | 0.01 (0.95) |
| 12mo (n = 25) | Fluency | -0.62 (0.001)* | 0.09 (0.66) | 0.07 (0.75) | 0.11 (0.61) | 0.02 (0.92) |
|  | Syntactic accuracy | -0.76 (0.00001)* | 0.30 (0.14) | 0.06 (0.79) | 0.07 (0.73) | 0.14 (0.52) |
|  | Lexical selection | -0.38 (0.06) | -0.34 (0.10) | -0.13 (0.55) | 0.31 (0.14) | 0.41 (0.04)^#^ |
|  | Structural complexity | -0.57 (0.003)* | 0.07 (0.73) | 0.10 (0.66) | 0.11 (0.59) | 0.19 (0.36) |

Numbers outside/inside parentheses denote the r/*P* values. *: Bonferroni correct *P* < 0.05/5; #: *P* < 0.05.

**SI Table 4. Lesion-symptom mapping results of white matter tract damage associated with decreased recovery in four aspects of connected speech from acute to three time points in the year following stroke.**

|  |  | ASAF | LSAF | PSAF | FAT | IFOF | ILF | UF |
| --- | --- | --- | --- | --- | --- | --- | --- | --- |
| subacute (n = 31) | Fluency | 0.70 (0.24) | 0.75 (0.23) | -0.42 (0.67) | 3.34 (0.001)***** | -0.36 (0.64) | -0.53 (0.71) | -0.62 (0.73) |
|  | Syntactic accuracy | 0.11 (0.45) | 0.94 (0.17) | 0.33 (0.37) | 1.91 (0.03)**^#^** | -0.69 (0.75) | -0.23 (0.59) | -1.46 (0.90) |
|  | Lexical selection | -0.33 (0.63) | 0.83 (0.20) | 0.86 (0.19) | -1.81 (0.96) | -0.43 (0.67) | 0.09 (0.46) | 1.93 (0.06) |
|  | Structural complexity | 0.14 (0.44) | 1.02 (0.16) | 0.47 (0.32) | -0.61 (0.72) | -1.37 (0.91) | -1.55 (0.93) | -0.39 (0.65) |
| 6mo (n = 31) | Fluency | -0.73 (0.77) | -0.42 (0.66) | -1.55 (0.92) | 0.49 (0.31) | -0.35 (0.64) | -0.90 (0.82) | -0.53 (0.70) |
|  | Syntactic accuracy | 0.60 (0.27) | 0.32 (0.37) | 2.91 (0.01)**^#^** | -0.96 (0.83) | -0.51 (0.69) | 0.89 (0.19) | -1.10 (0.86) |
|  | Lexical selection | 0.39 (0.35) | 0.77 (0.23) | 0.93 (0.18) | 0.04 (0.49) | 0.76 (0.23) | 1.20 (0.12) | -0.58 (0.72) |
|  | Structural complexity | -0.83 (0.80) | 0.08 (0.47) | -1.86 (0.95) | 2.72 (0.006)***** | -0.19 (0.58) | -0.35 (0.64) | -0.66 (0.75) |
| 12mo (n = 25) | Fluency | 1.10 (0.15) | 0.92 (0.18) | 0.69 (0.25) | -0.40 (0.66) | -1.42 (0.92) | -0.66 (0.75) | -1.18 (0.87) |
|  | Syntactic accuracy | 1.08 (0.15) | 2.73 (0.01)**^#^** | 0.72 (0.24) | -0.35 (0.65) | -1.43 (0.92) | 0.31 (0.38) | -3.31 (0.98) |
|  | Lexical selection | -0.37 (0.65) | -0.41 (0.66) | 0.00 (0.51) | 0.46 (0.32) | 0.48 (0.31) | -0.35 (0.64) | 0.82 (0.21) |
|  | Structural complexity | -1.31 (0.89) | -0.49 (0.70) | -3.34 (0.99) | 3.20 (0.007)***** | -0.23 (0.59) | -1.14 (0.86) | 0.88 (0.20) |

Numbers outside/inside parentheses denote the z/*P* values. *: Bonferroni correct *P* < 0.05/7; #: *P* < 0.05. ASAF: anterior segment of arcuate fasciculus; LSAF: long segment of arcuate fasciculus; PSAF: posterior segment of arcuate fasciculus; FAT: frontal aslant tract; IFOF: inferior fronto-occipital fasciculus; ILF: inferior longitudinal fasciculus; UF: uncinate fasciculus.

**SI Table 5. Lesion-symptom mapping results of white matter tract damage associated with decreased recovery in four aspects of connected speech for patients tested at acutely, and all the three time points in the year following stroke (subacute, 6 months, 12 months; n= 20).**

|  |  | ASAF | LSAF | PSAF | FAT | IFOF | ILF | UF |
| --- | --- | --- | --- | --- | --- | --- | --- | --- |
| subacute (n = 20) | Fluency | -0.57 (0.71) | -0.57 (0.72) | -0.40 (0.66) | 0.88 (0.19) | -1.33 (0.90) | -0.88 (0.81) | -0.66 (0.75) |
|  | Syntactic accuracy | -0.54 (0.71) | 0.66 (0.25) | 0.32 (0.37) | 0.79 (0.21) | -1.57 (0.93) | -0.36 (0.65) | -1.86 (0.92) |
|  | Lexical selection | 0.93 (0.18) | 1.59 (0.07) | 1.71 (0.07) | -0.67 (0.76) | 1.47 (0.08) | 2.23 (0.05) **^#^** | 3.03 (0.04) **^#^** |
|  | Structural complexity | 0.30 (0.38) | 0.95 (0.17) | 0.46 (0.32) | 0.40 (0.35) | -0.91 (0.82) | -1.40 (0.89) | -2.66 (0.95) |
| 6mo (n = 20) | Fluency | -0.93 (0.82) | -0.37 (0.65) | -0.31 (0.62) | -0.78 (0.78) | -1.21 (0.88) | -2.16 (0.95) | -0.28 (0.62) |
|  | Syntactic accuracy | 0.68 (0.24) | 0.43 (0.34) | 2.05 (0.05) | -0.24 (0.59) | 0.08 (0.46) | 0.37 (0.35) | -0.31 (0.62) |
|  | Lexical selection | 1.55 (0.09) | 1.46 (0.09) | 1.20 (0.13) | -0.61 (0.73) | 4.42 (0.003)* | 2.25 (0.04) **^#^** | 2.42 (0.06) |
|  | Structural complexity | -0.15 (0.56) | 0.21 (0.41) | -1.40 (0.89) | 2.46 (0.02)**^#^** | 0.33 (0.37) | 0.44 (0.32) | -0.97 (0.81) |
| 12mo (n = 20) | Fluency | 0.85 (0.20) | 0.77 (0.22) | 0.57 (0.28) | -0.11 (0.55) | -1.40 (0.91) | -1.13 (0.86) | -0.68 (0.75) |
|  | Syntactic accuracy | 1.16 (0.14) | 2.54 (0.02)**^#^** | 1.02 (0.16) | 0.04 (0.49) | -0.71 (0.76) | 0.48 (0.31) | -1.68 (0.90) |
|  | Lexical selection | 0.17 (0.43) | 0.16 (0.44) | 0.55 (0.29) | -0.28 (0.62) | 0.27 (0.39) | 0.20 (0.42) | -0.23 (0.60) |
|  | Structural complexity | -1.04 (0.84) | -0.52 (0.70) | -2.94 (0.98) | 2.14 (0.03) **^#^** | -0.32 (0.63) | -0.76 (0.78) | 0.34 (0.37) |

Numbers outside/inside parentheses denote the z*/P* values. *: Bonferroni correct *P* < 0.05/7; #: *P* < 0.05. ASAF: anterior segment of arcuate fasciculus; LSAF: long segment of arcuate fasciculus; PSAF: posterior segment of arcuate fasciculus; FAT: frontal aslant tract; IFOF: inferior fronto-occipital fasciculus; ILF: inferior longitudinal fasciculus; UF: uncinate fasciculus.

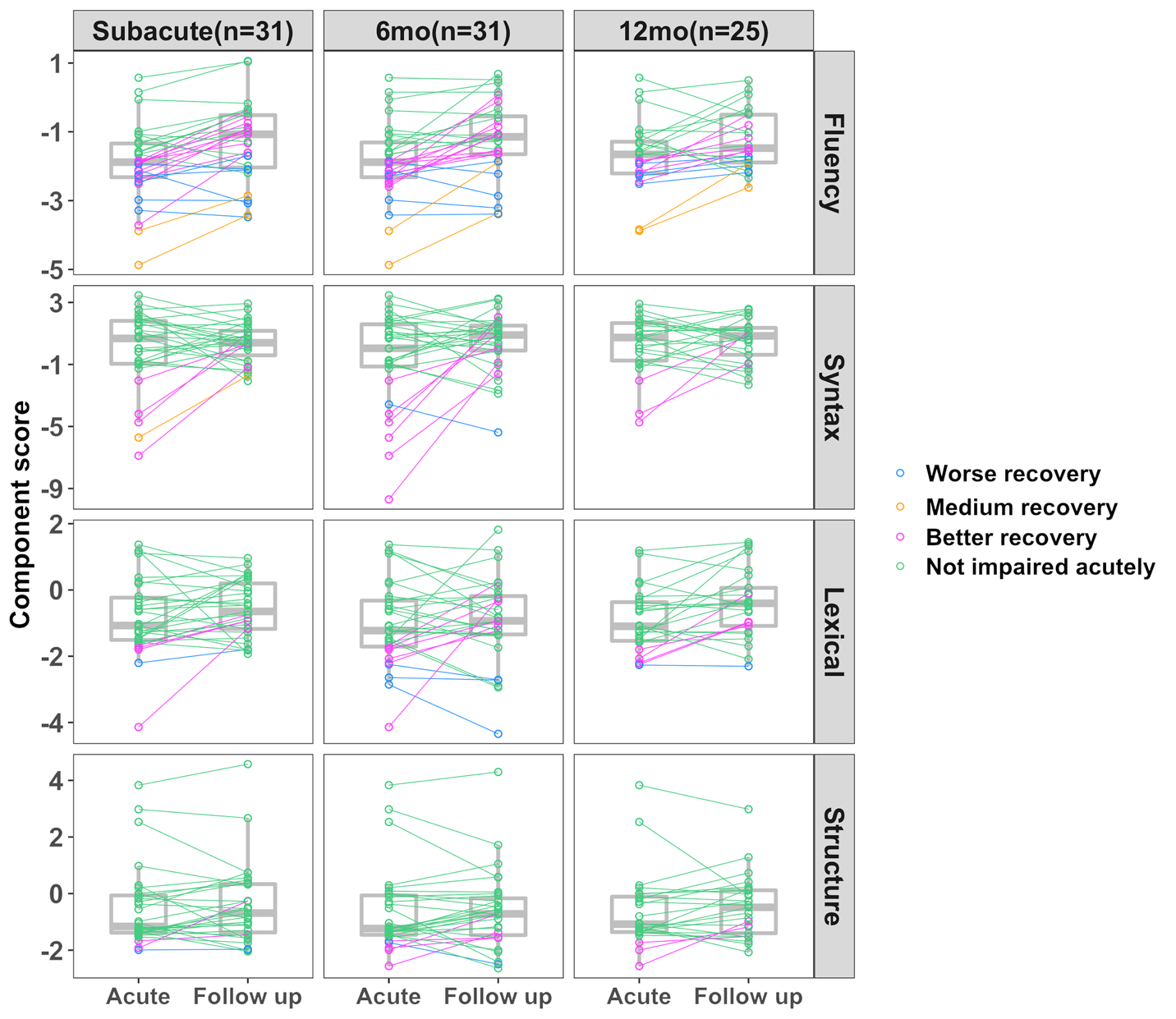

**SI Figure 1. Different recovery trajectories of four aspects of connected speech from acute to three follow-up time points in the year following stroke.** Worse recovery was defined as both acute performance worse than 95% of controls’ performance (z’s < -1.67) and follow up performance still worse than 95% of controls (z’s < -1.67; in blue). Medium recovery was defined as acute performance worse than 95% of controls’ performance (z’s < -1.67) where large gains occurred (recovery > 1 SD) but not to within the control range (in yellow). In contrast, better recovery was defined as those with impaired acute performance z’s < -1.67 but subsequent follow up performance was better than 95% of control performance (z’s > -1.67; in magenta). We defined not impaired as individuals who performed better than 95% of controls’ performance acutely (z’s > -1.67; in green).

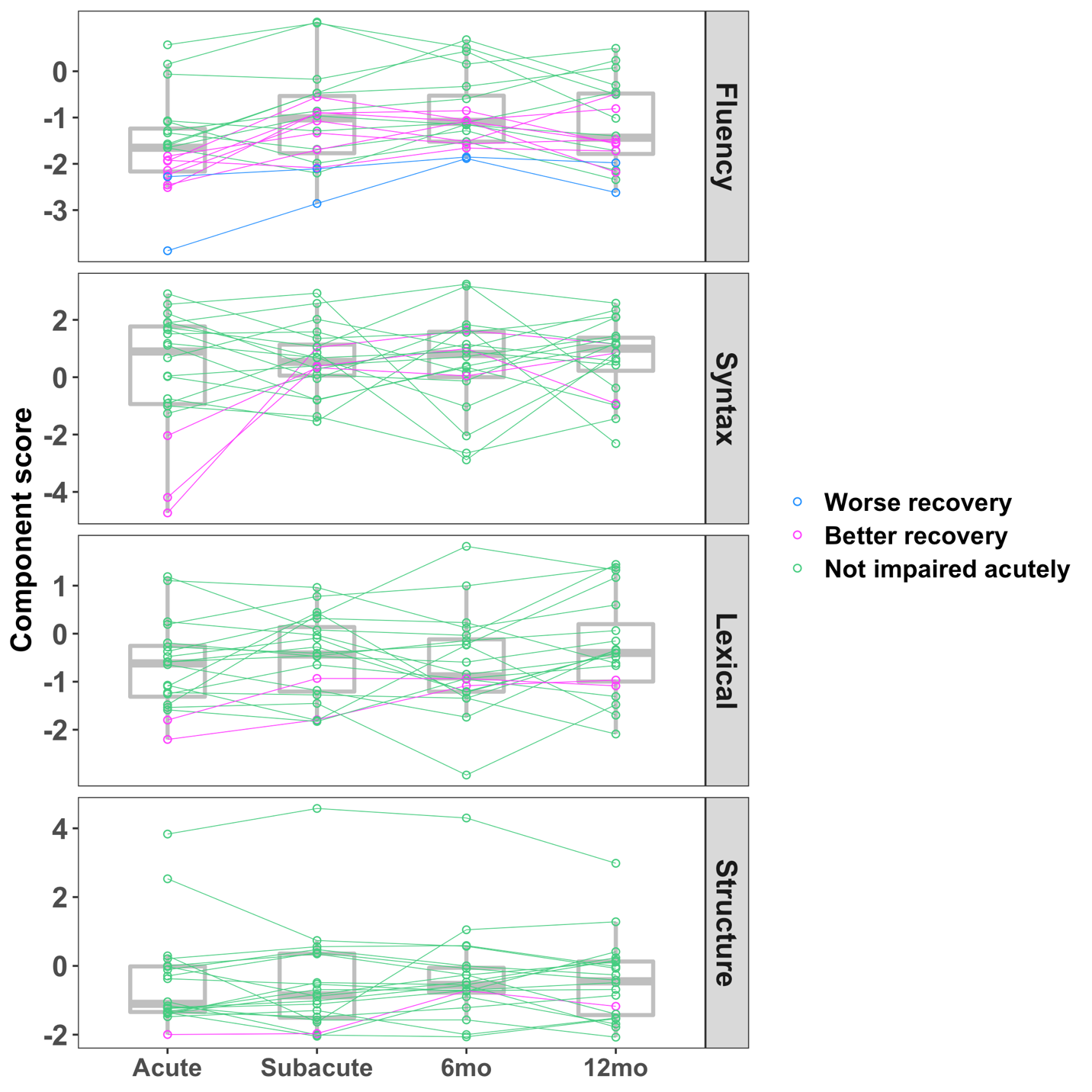

**SI Figure 2. Recovery of four aspects of connected speech for the 20 patients testing from acute to three follow-up time points in the year following stroke.** Worse recovery was defined as both acute performance worse than 95% of controls’ performance (z’s < -1.67) and follow up performance still worse than 95% of controls (z’s < -1.67; in blue). In contrast, better recovery was defined as those with impaired acute performance z’s < -1.67 but subsequent follow up performance better than 95% of control performance (z’s > -1.67; in magenta). We defined not impaired as individuals who performed better than 95% of controls’ performance acutely (z’s > -1.67; in green).

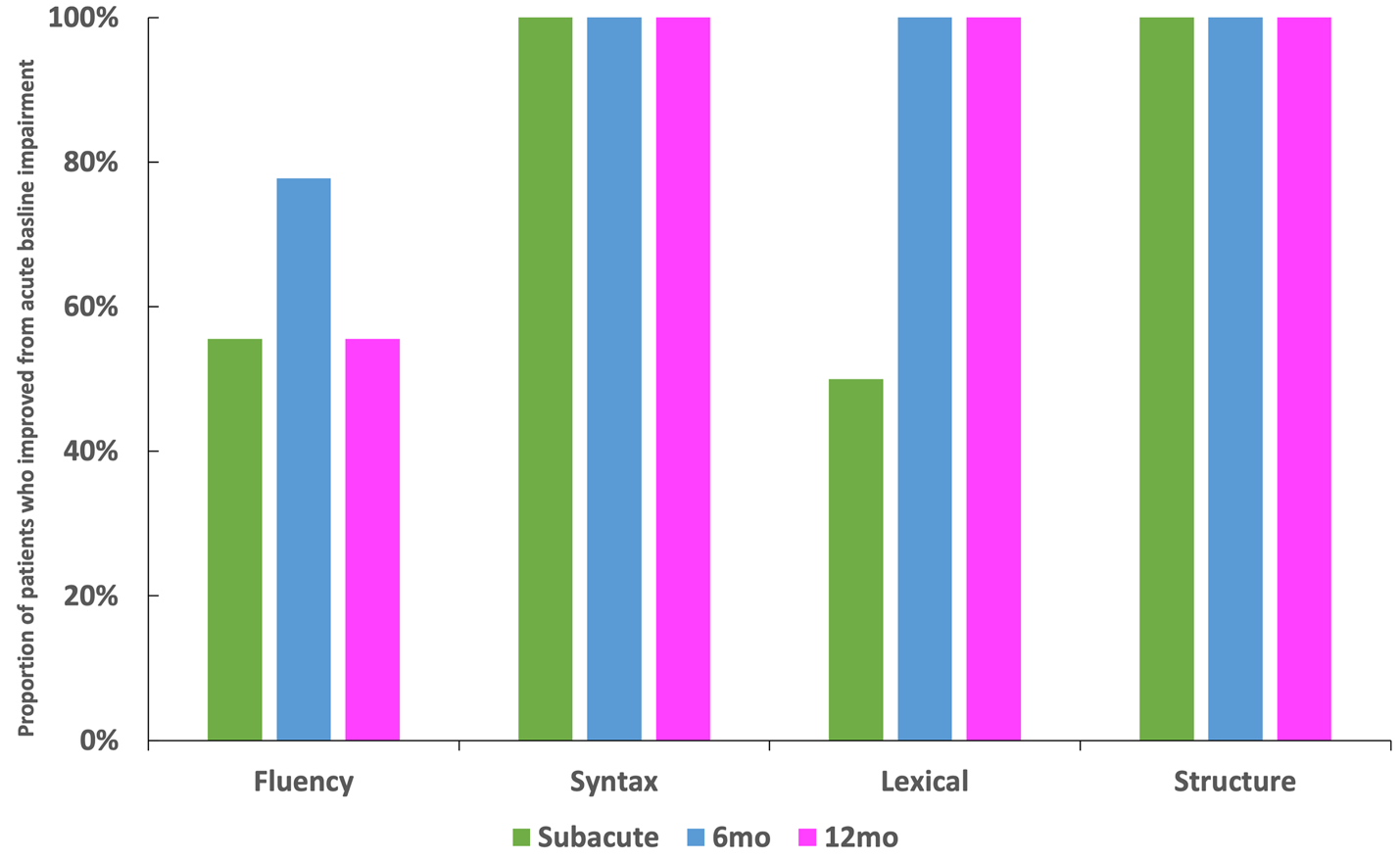

**SI Figure 3. The proportions of the 20 patients testing at all the timepoints who recovered to within normal levels (z > -1.67) after beginning with impairment acutely (z < -1.67) in four aspects of connected speech across three time points in the year following stroke.**

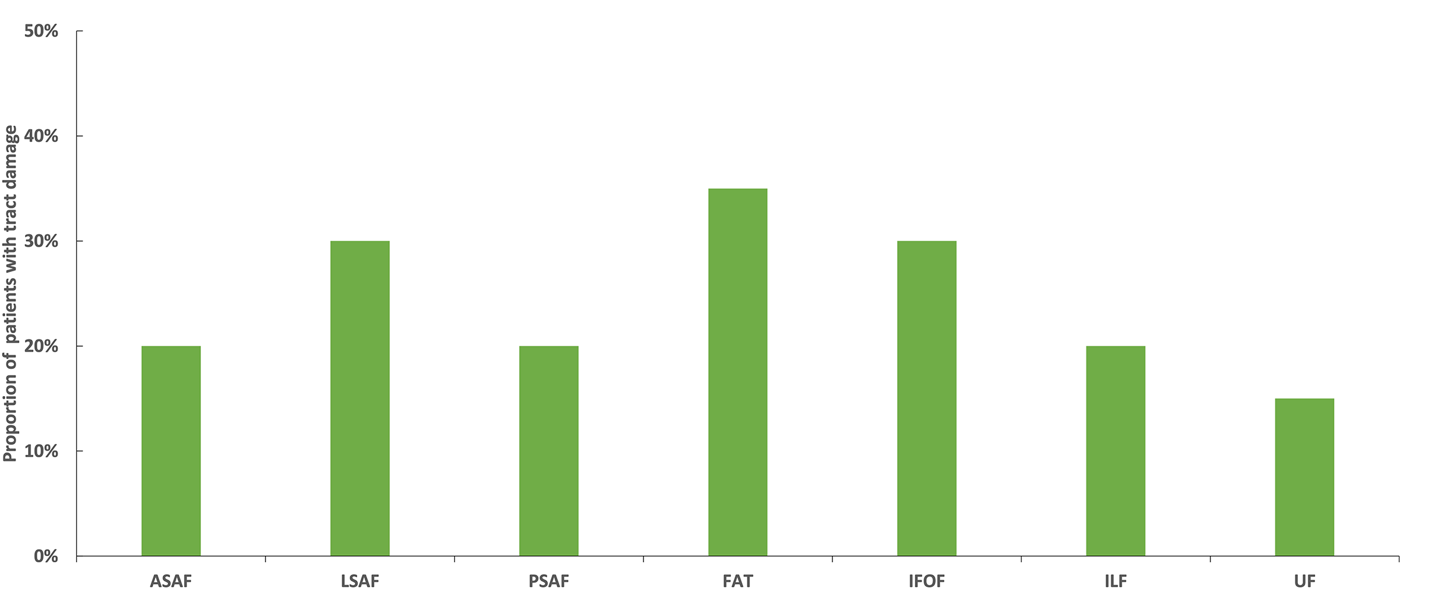

**SI Figure 4. Proportion patients with tract damage for each tract in the patient cohort testing at all the subacute, early chronic (6mo) and late chronic (12mo) time points (n = 20).** ASAF: anterior segment of arcuate fasciculus; LSAF: long segment of arcuate fasciculus; PSAF: posterior segment of arcuate fasciculus; FAT: frontal aslant tract; IFOF: inferior fronto-occipital fasciculus; ILF: inferior longitudinal fasciculus; UF: uncinate fasciculus.

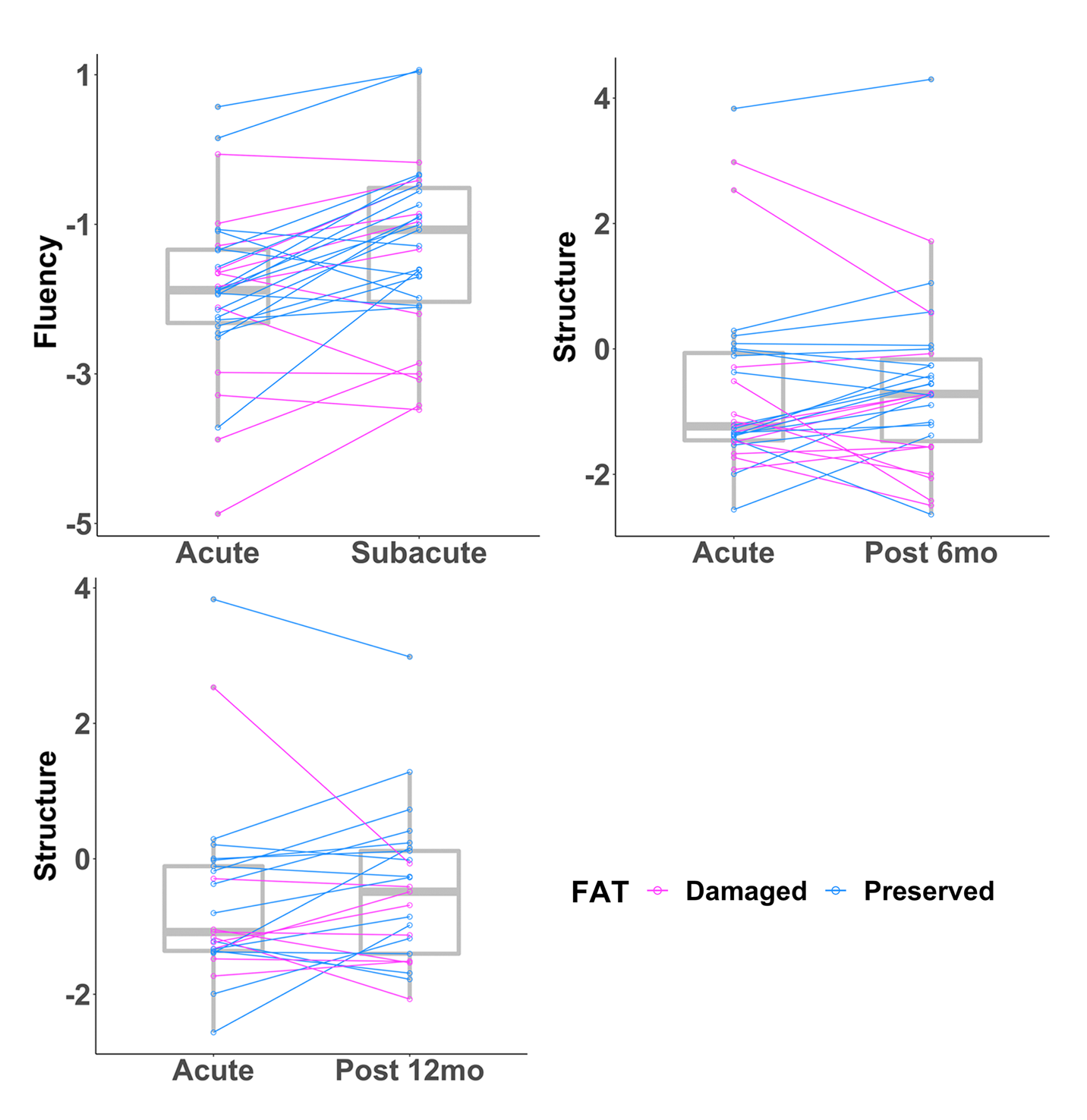

**SI Figure 5. The white matter tracts whose acute damage significantly predicted decreased recovery in four aspects of connected speech across three time points in the year following stroke (subacute, 6mo, and 12mo post stroke) without correction for multiple comparisons.** The box plots show the acute and follow-up connected speech scores clustered by tract damage. Magenta lines indicate subjects with damaged tracts, while blue lines indicate subjects with spared tracts. LSAF: long segment of arcuate fasciculus; PSAF: posterior segment of arcuate fasciculus; FAT: frontal aslant tract.
